# Food Colorings in Child-Targeted Ultra-Processed Foods in Brazil: Market Prevalence and Parental Perceptions

**DOI:** 10.64898/2026.06.12.26355247

**Authors:** Beatriz Silva Nunes, Maria Fernanda Pereira Eberle, Mariana F. Grilo, Camila Zancheta, Allison C. Sylvetsky, Ana Clara Duran

## Abstract

**Objective:** To assess the prevalence of food colorings in child-targeted ultra-processed foods in Brazil and explore parents’ perceptions of and ability to identify these additives.

**Design:** Mixed-methods study.

**Setting:** Brazil.

**Participants:** Quantitative data included 5,078 ultra-processed foods (UPFs) launched in Brazil between 2018 and 2021 from the Mintel Global New Products Database. Qualitative data came from seven focus groups with parents of children aged 2–5 and 6–11 years in Brazil.

**Phenomenon of Interest and Variables:** Presence of food colorings in child-targeted UPFs in Brazil and parents’ perceptions and identification of food colorings on food labels.

**Analysis:** Descriptive analyses and Thematic Framework Analysis.

**Results:** Of 5,078 UPFs, 23.0% had child-targeted marketing, and 40.3% of these contained food colorings. Prevalence was highest in carbonated beverages (88.0%), candies (73.7%), and ice cream (63.2%). Parents understood that food colorings make products attractive to children and considered them harmful, but said cost, availability, and children’s preferences made them hard to avoid. Most parents identified food colorings in ingredient lists, but many also classified other ingredients as colorings.

**Conclusions and Implications:** Food colorings are common in child-targeted UPFs in Brazil. Restricting child-directed marketing, labeling food colorings more clearly, and helping parents read ingredient lists could reduce children’s exposure.(204)

## 1. INTRODUCTION

Marketing strategies targeting children, such as packaging advertisements and the use of appealing colors, flavors, and shapes, are widely employed to increase product attractiveness and influence family purchasing decisions ^1^. Beyond packaging, children are also exposed to these strategies through television and social media ^1^, where advertising frequently promotes ultra-processed foods (UPFs) high in fats, sodium, sugars, and/or food additives ^2^. These practices are commercially effective but promote unhealthy dietary patterns and may harm children’s health ^3^.

In 2014, the Brazilian National Council for the Rights of Children and Adolescents (CONANDA) issued a resolution stating that advertising directed at children to induce consumption is illegal when it uses elements such as childish language, excessive colors, child characters, or promotional gifts ^4^. The food industry, however, did not recognize this resolution as having regulatory power ^5^, arguing that only federal legislation could legitimately ban advertising aimed at children. As a result, children remain exposed to persuasive tactics ^6^. Children are particularly vulnerable to marketing given their limited capacity to critically interpret marketing and the formative nature of early dietary habits, which may have lasting health implications ^7^.

UPFs can be identified by ingredients listed on food labels, including substances rarely used in culinary preparations and food additives with cosmetic functions ^2^. Food additives with cosmetic functions include food colorings, flavorings, sweeteners, and other substances used to modify sensory characteristics, such as adding or restoring color to foods ^2^. Among them, certain synthetic food colorings, such as Tartrazine, Brilliant Blue FCF, and Sunset Yellow FCF, have been linked to adverse health outcomes in children, including hyperactivity, neurotoxicity, and potential carcinogenic effects ^8–11^.

In Brazil, food additives are regulated by the National Health Surveillance Agency (ANVISA), which permits only substances that have been scientifically evaluated and approved for use ^12^. These regulations are broadly aligned with international standards, such as those from the Codex Alimentarius and the U.S. Food and Drug Administration (FDA) ^13^. Although this framework is designed to protect consumers, regulatory approvals are often based on the evidence available at the time, which may reflect limited consideration of the adverse effects associated with prolonged exposure during critical stages of development ^8^.

Monitoring packaged foods marketed to children is needed, particularly for ingredients linked to potential health risks, such as food colorings. Given the central role of parents in shaping children’s food choices, it is also important to understand how they perceive food additives and whether they can identify them on food labels ^14^. Such evidence can support the development and strengthening of regulations aimed at protecting the health rights of children ^15^.

Parents’ decisions about food colorings can be understood through the Theory of Planned Behavior (TPB), which proposes that behavior follows from intention, shaped by attitudes toward the behavior, subjective norms (perceived social pressure), and perceived behavioral control (the perceived ease or difficulty of performing the behavior, which depends on resources such as time and money) ^16,17^. Under this framework, parents’ attitudes toward food colorings, the norms they perceive, and the barriers they face, such as cost and the products available to them, may shape both their intention to avoid these additives and their ability to do so. Examining the products available on the market alongside parents’ perceptions can therefore identify key barriers to avoiding food colorings that parents encounter when making food and beverage choices for their children.

To our knowledge, no study in Brazil has examined food colorings specifically in products with child-targeted marketing or explored how parents perceive these additives and identify them on labels. This study aimed to (i) assess the prevalence of food colorings in foods and beverages with child-targeted marketing in the Brazilian food supply and (ii) explore parents’ perceptions of food colorings in beverages commonly consumed by children, including their ability to identify these additives on the package. The results can inform nutrition education for parents and regulatory policy aimed at reducing children’s exposure to child-directed marketing and food colorings.

## 2. METHODS

This mixed-methods study had 2 components: (1) a cross-sectional analysis of the Brazilian packaged food supply to estimate the prevalence of child-targeted marketing and food colorings, and (2) focus groups with parents followed by a brief questionnaire on the identification of food colorings on beverage labels.

### 2.1. Prevalence of Marketing Strategies and Food Colorings n Child-Targeted Ultra-processed Foods in Brazil

#### 2.1.1. Identification of ultra-Processed Foods

We used data from the Mintel Global New Products Database (GNPD) ^18^. This commercial database (hereinafter referred to as Mintel GNPD) monitors the launch of new food and beverage products in retail markets in more than 80 countries worldwide ^19^. These data have been widely used in nutrition-related research ^20,21^. In Brazil, Mintel GNPD data on nutritional composition and nutrition claims have been previously validated and shown to be suitable for evaluating public food labelling policies ^22,23^.

In this study, we used data on products launched in Brazil and recorded in the Mintel GNPD from 2018 to 2021 to assess the prevalence of food colorings in foods and beverages with child-targeted marketing in Brazil. We included only top-selling brands, representing 80% of the market share in each food category, as well as private-label brands from major Brazilian retailers, according to Euromonitor International, including Casino, Carrefour, WMB, Supermercados Cencosud, Supermercados BH, Cia Zaffari, and Supermercado Dia ^24^. The initial sample comprised 7,202 products, of which 5,599 were from top-selling brands and 1,603 were private-label products. We excluded products with illegible images or imported items (n = 288), those with incorrect sample year or barcode information (n = 157), and infant formulas (n = 42), resulting in a final sample of 6,715 products. Given the aim of our study to identify the presence of food colorings, our analysis focused solely on UPFs, which were identified based on the presence of markers in the ingredient list, following the method proposed by Grilo et al. ^25^. After excluding 1,637 products not classified as UPFs, the analytic sample comprised 5,078 UPFs.

#### 2.1.2. Identification of UPFs with child-targeted marketing

We identified products with marketing strategies targeting children by searching for child-directed elements on the front of the packaging label, using a protocol developed by Borges et al. (under review). Five trained nutritionists coded these strategies using a validated protocol based on the INFORMAS framework ^26^ and the World Health Organization food label monitoring protocols ^27^, as detailed by Kikuta et al. ^22^. The reliability of data collection was evaluated through intra- and inter-rater analyses on 10% of the sample, using duplicate data entries by the same individual and two different individuals, with high agreement (κ > 0.90).

The child-targeted marketing elements included references to unconventional effects generated by consumption (e.g., explodes in the mouth, gives you superpowers), fun, cool, or adventurous themes (e.g., full of adventures, super cool), and products with unconventional shapes (e.g., star-shaped, teddy bear-shaped, or car-shaped items). We also included brand mascots, licensed characters, references to sports in general (e.g., images of balls, football, or basketball), references to professional athletes and/or famous sports teams, references to famous sporting events (e.g., the FIFA World Cup, the Olympic Games), school-related items, and the inclusion of gifts or prizes (Borges et al. under review). We excluded references to taste, smell, color, or texture, as such features may also appear on products not specifically targeted at children (e.g., mayonnaise or bouillon cubes).

#### 2.1.3. Identification of food colorings

To identify the 51 food colorings listed in the Codex Alimentarius General Standard for Food Additives ^28^, we searched the list of ingredients for their names, synonyms, or INS numbers (e.g., Fast Green FCF, or INS 134, or C.I. Food Green 3). Some food additives, such as caramels, annatto, iron oxides, and lycopene, were grouped due to the non-specific way they are often labeled; for example, we classified both “synthetic lycopene” and generic declarations of “lycopene” within the same lycopene group ^29^. In addition, we excluded food colorings that may be added as vitamins and minerals in foods, including food additives from the beta-carotene and riboflavin groups, as well as calcium carbonate. Full details of these groupings are provided in Supplementary Material 1.

#### 2.1.4. Food categories

We classified the selected UPFs into 14 categories: cured meats; sweet biscuits; savory biscuits; cakes and pies; bread; candies in general; carbonated beverages; chocolate; ready-to-eat meals; non-carbonated beverages; dairy beverages; ice cream; sauces and condiments; and other UPFs, based on the classification of the 2017-2018 Brazilian Household Budget Survey ^30^.

### 2.2. Focus Groups to Examine Parents’ Perceptions of Food Coloring in Beverages Commonly Consumed by Children

The qualitative component was conducted within a broader study examining parents’ perceptions of non-sugar sweeteners in beverages consumed by children [blinded for review], which included questions on parents’ perceptions of food colorings. Given that the focus group methodology and findings pertaining to non-sugar sweeteners were published previously, we briefly describe the procedures specific to food colorings in the sections below.

Seven focus groups (3–8 participants each) were conducted in person with 40 parents between July and November 2023 in urban areas of two cities of state of São Paulo, Brazil. Groups were also stratified by the type of school the child attended (public or private), as a proxy for socioeconomic position. Inclusion criteria were being a parent or caregiver of a child aged 2–11 years, living in the same household as the child, and being responsible for at least half of the household’s grocery or food shopping. Parents of children with diet-related conditions requiring special diets were excluded, as they tend to be more attentive to nutrition labels ^32^.

Focus group discussions were conducted separately with parents of children aged 2–5 years and parents of children aged 6–11 years due to differences in beverage exposure and consumption in these age ranges ^33,34^. Prior to the focus group discussions, parents completed a brief sociodemographic questionnaire (Supplementary Material 2). After the focus group discussions, parents completed a questionnaire (Supplementary Material 3) assessing their ability to identify food colorings on product labels. We focused specifically on beverages because they represent an important source of food additives in children’s diets (Grilo et al., 2022).

#### 2.2.1 Procedures

Focus group discussions were led by a trained nutritionist with experience in qualitative research, supported by a research assistant, using a semi-structured discussion guide developed by the research team (Supplementary Material 4). All discussions were audio-recorded with participants’ consent. The questions were adapted from previous research on public perceptions of food additives and modified to include specific prompts related to food colorings ^36^.

During the discussions, participants were asked to reflect on the types of beverages they provide to their children and the main factors influencing their choices, including nutritional considerations, taste preferences, marketing, and affordability. This initial discussion helped contextualize participants’ decision-making processes regarding beverage selection. Subsequently, participants were asked about their perceptions of the function of food colorings, their children’s consumption of these additives, and their views on the potential health implications associated with food colorings.

After the focus group discussions, participants completed a questionnaire developed by the research team, which assessed their familiarity with identifying food additives, including food colorings, based on product labels. Parents were shown 5 beverages (flavored yogurt, milk, chocolate milk, soda, and powdered juice) and asked to indicate which contained food colorings, first based only on the front of the package and then by marking the food colorings in each ingredient list. These beverages are widely consumed by children in Brazil, and the brands shown were the top sellers in each category according to Euromonitor market share data ^37^. For the present study, only questions related to food colorings were analyzed.

### 2.3. Statistical Analysis

#### 2.3.1. Prevalence of marketing strategies and food colorings in child-targeted UPFs in Brazil

We conducted descriptive analyses with 95% confidence intervals (CIs) to estimate the prevalence of child-targeted marketing strategies in UPFs and the prevalence of food colorings among child-targeted UPFs, both overall and by food category. We also identified the 10 most frequently used food colorings among child-targeted UPFs.

#### 2.3.2. Focus groups to examine parents’ perceptions of food coloring in beverages commonly consumed by children

As described previously (blinded for review), audio recordings of the focus group discussions were transcribed and translated bi-directionally (Portuguese to English and back) by a native speaker, with two bilingual reviewers ensuring accuracy. Transcripts were then independently coded by two trained researchers using Dedoose™ software. Researchers followed a structured analysis approach, including familiarization with the data, application of deductive codes, and iterative identification of inductive themes. A codebook was developed and refined to ensure consistency. A deductive-inductive coding approach was applied; deductive coding addressed predefined research questions, while inductive coding allowed new themes to emerge ^38^. Thematic Framework Analysis guided the process to ensure both expected and emergent insights were captured ^39^.

Discrepancies were resolved through discussion with a third researcher. Representative quotes were selected to illustrate key themes ^40^. To enhance objectivity, transparency, and rigor, the team held regular discussions, refined codes iteratively, and had all data independently coded by 2 researchers.

Questionnaire responses were summarized as percentages for each beverage: participants who classified the product as containing food colorings based on the front of the package, participants who correctly identified the food colorings in the ingredient list, and participants who classified other ingredients as food colorings.

Statistical analyses were conducted using Stata/MP 16.1 (StataCorp LLC) and qualitative analyses were performed using Dedoose™.

### 2.4. Ethical Aspects

This study received approval from the National Ethics Committee and the Research Ethics Committee of XXX (blinded for review). All participants provided informed consent before completing any questionnaires or beginning the focus group discussion.

## 3. Results

### 3.1. Prevalence of marketing strategies and food colorings in child-targeted UPFs in Brazil

Among the 5,078 UPFs analyzed in the Mintel GNPD, 1,168 products (23.0%; 95% CI: 21.8–24.1) featured at least one marketing strategy targeted at children. Among these products, a total of 1,616 marketing strategies were identified; the most prevalent were references to fun, cool, or adventurous themes (45.4%), unconventional product shapes (31.8%), and the presence of brand mascots (31.7%), as shown in Table 1.

**Table 1.** Marketing strategies targeting children among ultra-processed foods with child-targeted marketing (n = 1,168)

| **Child-targeted marketing strategies** | n | % (95% CI*) |
|---|---|---|
| Unconventional consumption effects | 33 | 2.8 (2.0 - 3.9) |
| References to fun, cool, or adventurous themes | 530 | 45.4 (42.5 - 48.2) |
| Unconventional product shapes | 371 | 31.8 (29.2 - 34.5) |
| Presence of brand mascots | 370 | 31.7 (29.1 - 34.4) |
| Presence of licensed characters | 174 | 14.9 (13.0 - 17.1) |
| References to sports (general) | 32 | 2.7 (1.9 - 3.8) |
| References to professional athletes and/or famous sports teams | 21 | 1.8 (1.2 - 2.7) |
| References to famous sporting events | 16 | 1.4 (0.8 - 2.2) |
| References to school-related themes or items | 29 | 2.5 (1.7 - 3.6) |
| Inclusion of gifts or prizes | 40 | 3.4 (2.5 - 4.6) |

Note: \*95% CI: 95% confidence interval. Products could display more than 1 strategy (1,616 strategies in total); percentages do not sum to 100.

Food colorings were present in 40.3% (95% CI: 37.5–43.2) of products with child-targeted marketing. The food categories with the highest prevalence of these food additives were carbonated beverages (88.0%; 95% CI: 68.7–96.1), candies (73.7%; 95% CI: 68.0–78.6), and ice creams (63.2%; 95% CI: 40.3–81.3) (Table 2).

**Table 2.** Prevalence of ultra-processed foods with child-targeted marketing containing food colorings in the Brazilian food market, by food category, between 2018 and 2021.

| **Food categories** | **Total** | **Products with food coloring** |  |
|---|---|---|---|
|  | **N** | **n** | **% (95% CI\*)** |
| Cured meats | 22 | 10 | 45.5 (26.5 - 65.9) |
| Sweet biscuits | 71 | 28 | 39.4 (28.8 - 51.2) |
| Savory biscuits | 82 | 37 | 45.1 (34.7 – 56.0) |
| Cakes and pies | 82 | 19 | 23.2 (15.3 - 33.5) |
| Bread | 64 | 1 | 1.6 (0.2 - 10.3) |
| Candies | 262 | 193 | 73.7 (68.0 - 78.6) |
| Carbonated beverages | 25 | 22 | 88.0 (68.7 - 96.1) |
| Chocolate | 314 | 69 | 22.0 (17.7 - 26.9) |
| Ready-to-eat meals | 19 | 8 | 42.1 (22.6 - 64.4) |
| Non-carbonated beverages | 59 | 11 | 18.6 (10.6 - 30.6) |
| Dairy drinks | 72 | 31 | 43.1 (32.2 - 54.7) |
| Ice cream | 19 | 12 | 63.2 (40.3 - 81.3) |
| Sauces and condiments | 38 | 19 | 50.0 (34.6 - 65.4) |
| Other Ultra-processed foods** | 39 | 11 | 28.2 (16.3 - 44.1) |
| Total | 1,168 | 471 | 40.3 (37.5 - 43.2) |

Note: \*95% CI: 95% Confidence Interval; \*\*Other UPFs are composed mainly of cheeses and cheese-based ultra-processed products, such as ricotta cream cheese and strawberry creamy quark.

In addition, the ten most prevalent food colorings in our sample are illustrated in Figure 1. Sunset Yellow FCF was the most frequent (16.9% of child-targeted UPFs), followed by Brilliant Blue FCF (14.7%) and Allura Red (14.3%). It is important to note that a single product may contain more than one food coloring.

**Figure 1.**
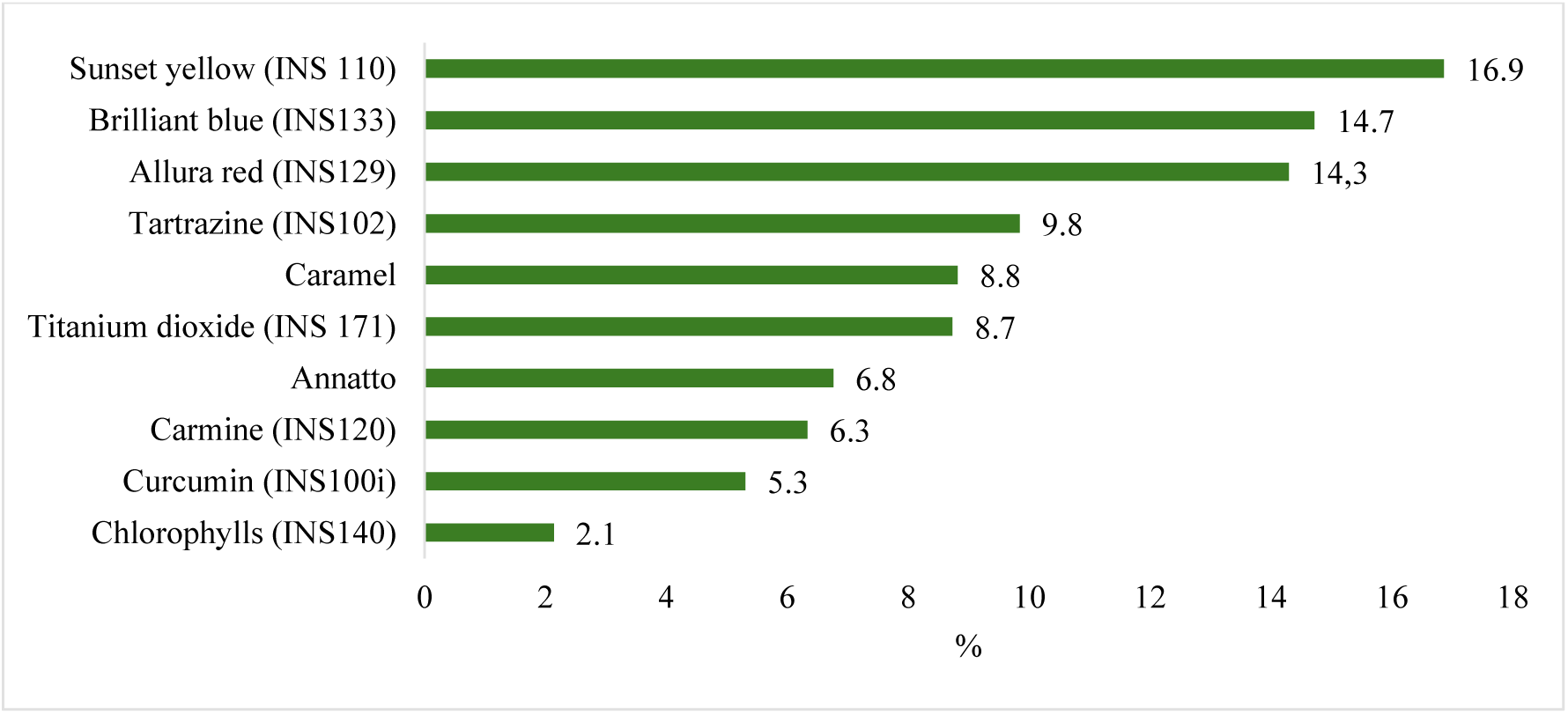
Prevalence (%) of the ten most common food colorings among ultra-processed foods with child-targeted marketing launched in the Brazilian food supply between 2018 and 2021 (n=1,168).

### 3.2. Parents’ Perceptions of Food Colorings in Beverages Commonly Consumed By Children

Most of the 40 participants were women (87,5%) and White (60%), and 60% had completed a college degree. Further details on the sociodemographic characteristics are provided in Supplementary Material 5.

Three main themes emerged from the focus group discussions: parents’ familiarity with the concept and origin of food colorings; their perception that these additives are unhealthy and their desire to avoid them; and their perception that products with food colorings enhance the visual appeal to children, particularly beverages. These themes are described in detail below.

The first theme reflected participants’ awareness of the term “food coloring.” Participants described food colorings as food additives used to give color to foods and beverages (e.g., “*It’s just something to add color […]”-* Group 3, public school), and explained that these food additives can be either artificial or natural (e.g. “*I’ve seen some labels that it can be natural or synthetic, right?” -* Group 5, public school). Participants were familiar with how food colorings are listed on product labels. However, they found this information confusing and incomplete, for example because it does not state the amount used (e.g., *“In the ingredients on the packaging, they come as symbols, you know, letters and numbers, that we don’t even know what they are. It should be better specified how much food coloring there is”* - Group 4, private school).

The second theme focused on health concerns and the desire to avoid food coloring, particularly artificial ones, in their children’s diets. Parents perceived food colorings as potentially harmful to children’s health and mentioned possible links to allergies and other long-term adverse effects (e.g., *“I think it’s terrible because they say it causes a lot of allergies”* Group 7, private school). This concern appeared to influence their purchasing decisions and attitudes toward certain products (Table 3).

**Table 3.**
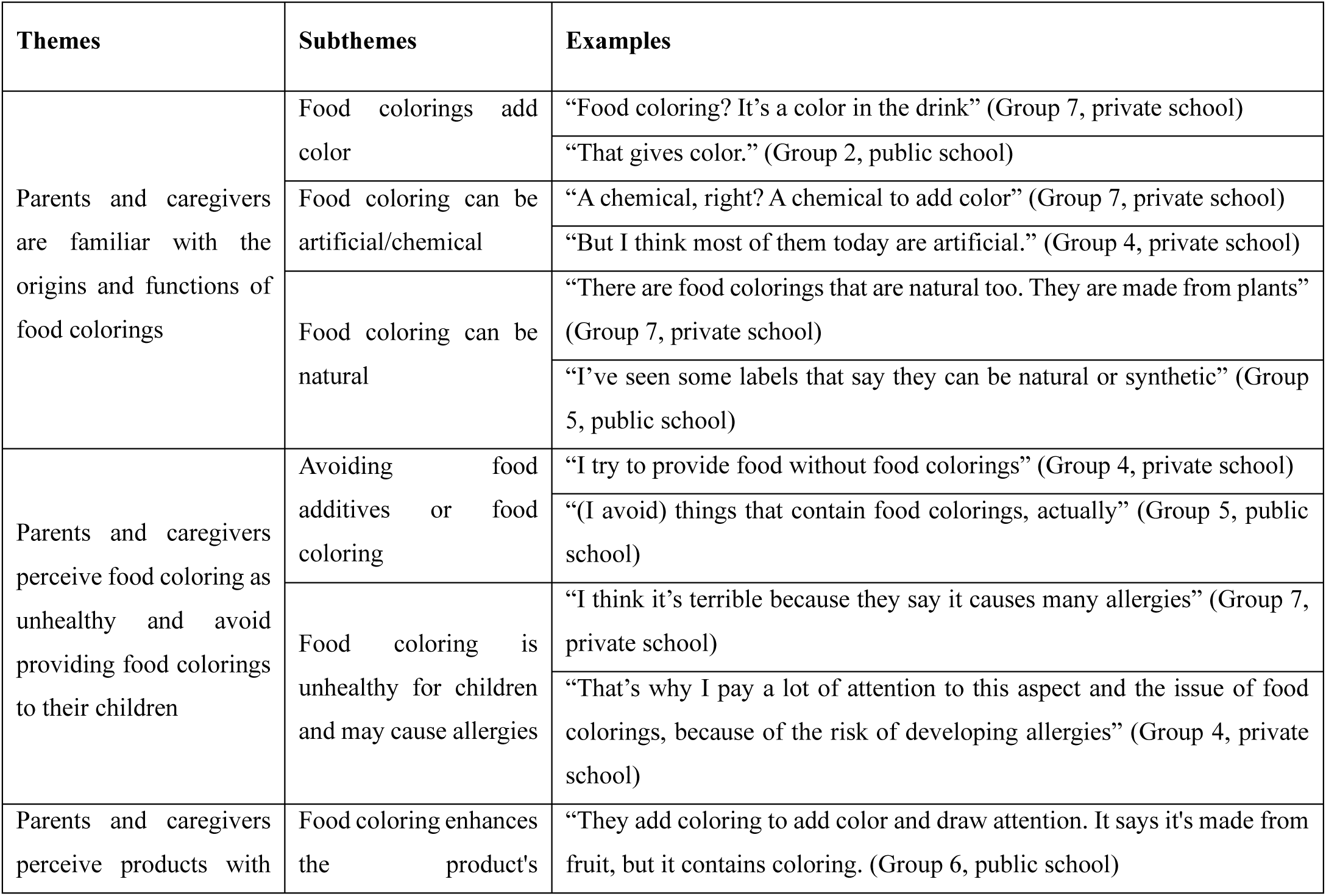
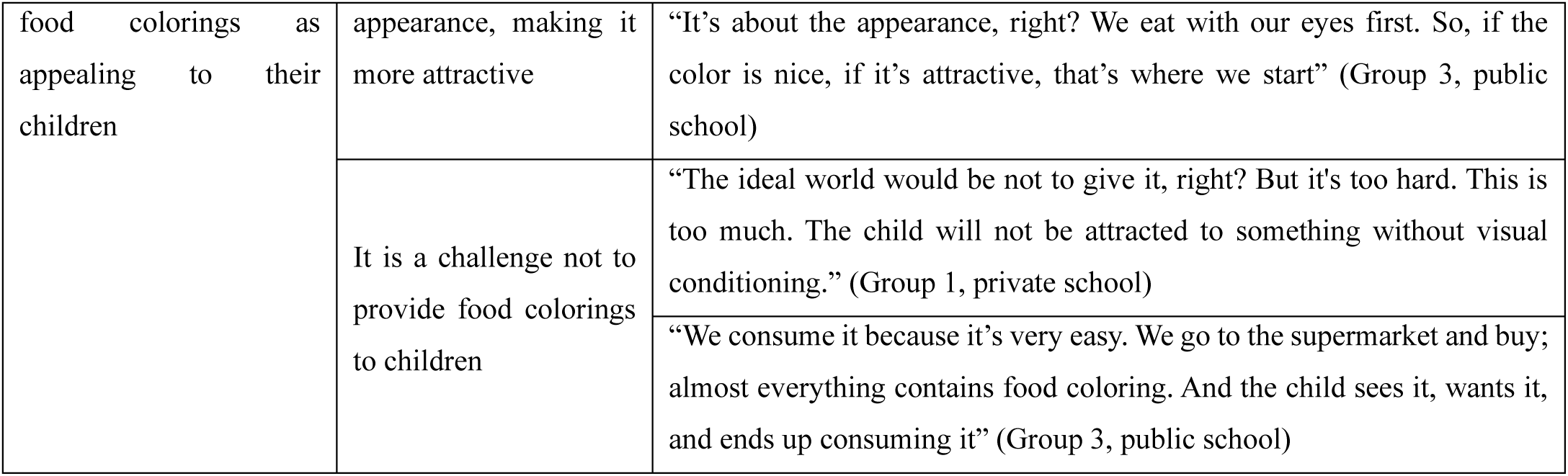
Themes and subthemes identified through focus groups.

However, in both public and private school groups, parents explained that, despite their intentions to avoid beverages with food colorings, they could not always do so because products without food colorings were often more expensive (e.g., *“I try to avoid it a lot, but that’s what we can offer, right, because natural juice is actually very expensive”* Group 5, public school; and *“This natural juice is one of the most expensive. The apple-only [fermented milk]. The orange juice. They are the most expensive, so it gets more difficult.” -* Group 4, private school). While all parents expressed some concern about food colorings, some viewed natural food colorings more favorably than artificial ones (e.g. *“If I recall correctly, there is an option of natural or synthetic food coloring; I’ve come across the natural one on the market. I make an effort to avoid the synthetic ones”* Group 5, public school) (Table 3).

The third theme focused on the role of food colorings in enhancing the visual appeal of products, especially beverages (e.g. *“It’s not good, but unfortunately, it’s an attraction. Nobody likes colorless drinks, right? It’s an attraction for children, it’s an attraction for humans, unfortunately”* - Group 7, private school). Participants discussed how specific food colorings are commonly associated with certain flavors, such as purple with grape and red with strawberries, and noted that many products rely on colorings and flavorings rather than whole ingredients (e.g., *“But just the color is understood. So, they want a strawberry pink, because that one over there (purple) will be grape, that’s what draws attention” -* Group 3, public school). Participants also explained that bright colors, along with other marketing strategies such as cartoon characters, capture children’s attention and contribute to their preference for these products. This combination of visual appeal and marketing was viewed as a challenge to efforts aimed at limiting children’s consumption of such items (Table 3).

Within this theme, participants reported believing that most of the products they usually offer to their children contain food colorings (e.g. *“We supply it daily because basically all the food we provide to children has coloring, mine drinks juice all the time” -* Group 5, public school). They also described products with food colorings as ubiquitous and easily accessible (e.g. *“[…] It’s very easy, right? We consume it because it is very easy, we go to the supermarket and buy it, almost everything contains food colorings”* Group 3, public school).

### 3.3. Identification of Food Colorings on Labels

Of the participants who completed the questionnaire, more than 85% were familiar with the five beverage types presented, and approximately one-third indicated that they regularly purchase these products (Supplementary Material 6).

Based on the front of the package, most participants correctly identified the beverages that contained food colorings (flavored yogurt, 94.9%; soda, 97.4%; powdered juice, 100.0%). However, 76.9% classified chocolate milk, which did not contain food colorings, as containing them, as did 15.4% for milk (Table 4). Using the ingredient lists, 79.5% to 89.7% of participants correctly identified the food colorings in the 3 beverages that contained them. At the same time, 7.7% to 53.9% misclassified other ingredients, such as “malt extract,” “strawberry preparation,” and “flavorings,” as food colorings.

**Table 4.**
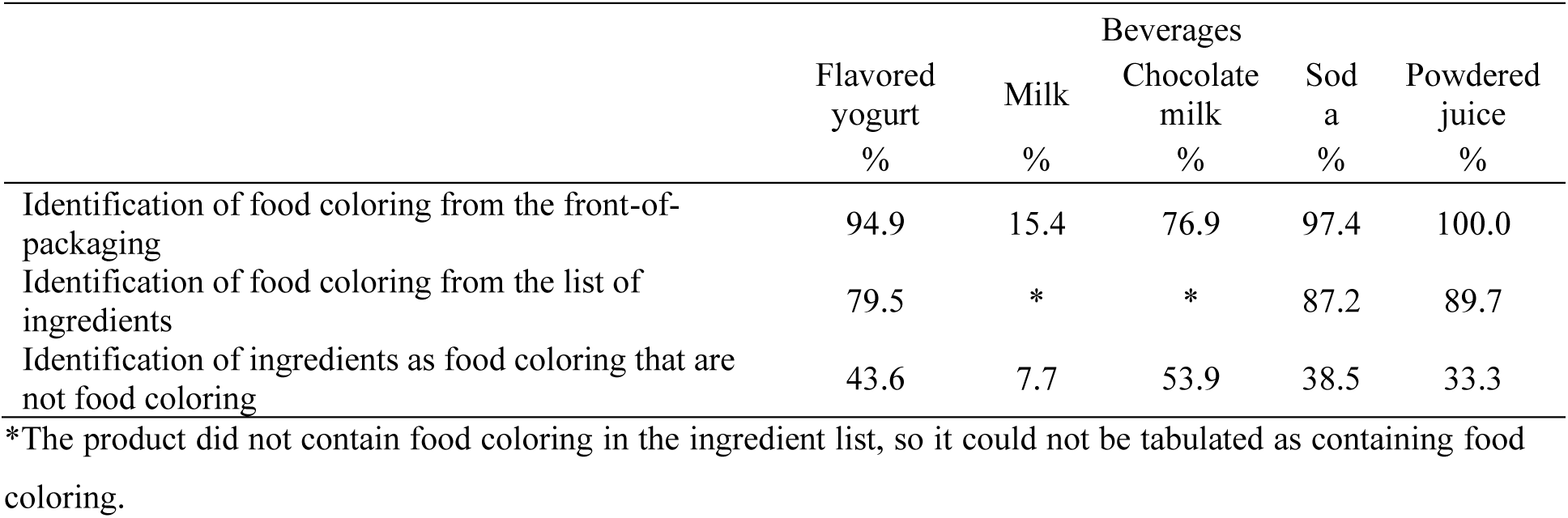
Identification of food coloring in the food labeling of five types of beverages by participants (n=40).

|  | Beverages |  |  |  |  |
|---|---|---|---|---|---|
|  | Flavored yogurt | Milk | Chocolate milk | Soda | Powdered juice |
|  | % | % | % | % | % |
| Identification of food coloring from the front-of-packaging | 94.9 | 15.4 | 76.9 | 97.4 | 100.0 |
| Identification of food coloring from the list of ingredients | 79.5 | * | * | 87.2 | 89.7 |
| Identification of ingredients as food coloring that are not food coloring | 43.6 | 7.7 | 53.9 | 38.5 | 33.3 |

\*The product did not contain food coloring in the ingredient list, so it could not be tabulated as containing food coloring.

## 4. DISCUSSION

This study provides novel evidence on the intersection of child-targeted marketing and food colorings in the Brazilian retail market. Approximately one-quarter of UPFs launched in the Brazilian retail market between 2018 and 2021 employed child-targeted marketing strategies, and 2 in 5 of these products contained food colorings, with Sunset Yellow FCF, Brilliant Blue FCF, and Allura Red being the most prevalent. Marketing strategies emphasizing fun, adventure, and playfulness were especially common. In the focus groups, parents understood the function of food colorings and viewed them as harmful, but described barriers to avoiding them: cost, the wide availability of products containing them, and the appeal of colorful, heavily marketed products to children. Most parents found food colorings in ingredient lists, but many also misclassified other ingredients and products as containing them.

These findings are consistent with previous Brazilian studies reporting widespread use of child-directed marketing in products with poor nutritional profiles, particularly candies, carbonated beverages, and other UPFs commonly consumed by children ^41^. In line with this literature, our results showed that carbonated beverages and candies had the highest prevalence of food colorings among products using child-targeted marketing strategies.

In 8 of the 14 food categories, more than 40% of child-targeted products contained food colorings: carbonated beverages, candies, ice cream, sauces and condiments, cured meats, savory biscuits, dairy drinks, and ready-to-eat meals. Many of these categories - particularly carbonated beverages, candies, and biscuits - are widely consumed by children and adolescents in Brazil ^42,43^,food coloring exposure may be widespread in this population. A similar pattern has been observed in Chile, where a study of preschool children’s exposure to food additives identified soft drinks and flavored yogurts as the main sources of additives in their diets ^44^.

Several of the most commonly identified food colorings in this study, such as Sunset Yellow, Brilliant Blue FCF, Allura Red, and Tartrazine, have been associated with adverse health outcomes, including hyperactivity, attention deficit, and hypersensitivity reactions such as urticaria, bronchospasm, rhinitis, angioedema, and even anaphylactic shock ^45^. Previous studies have also suggested that children may exceed the acceptable daily intake for additives such as Sunset Yellow FCF and others, given the increasing availability of these ingredients, highlighting the need to review the safety of food colorings already approved ^9–11^. The widespread presence of food colorings in products with child-targeted marketing is consistent with the themes that emerged from the focus groups in this study, in which participants perceived food colorings as ubiquitous and reported difficulties in avoiding products containing these additives in their children’s diets.

International regulatory agencies have increasingly adopted precautionary measures regarding these food colorings. In the United States, the FDA announced in 2025 a plan to phase out petroleum-based synthetic dyes, including Sunset Yellow, Brilliant Blue, Allura Red, and Tartrazine, from the food supply, largely through voluntary industry pledges tracked by the agency through 2027 ^46^, while in the European Union and the United Kingdom, products containing Sunset Yellow, Allura Red, Tartrazine, and 3 other azo dyes must carry a warning that they may have an adverse effect on activity and attention in children ^47,48^. In Brazil, all food additives used in foods and beverages must be declared in the ingredient list either by their full name or by their International Numbering System (INS) number ^49^. However, Brazilian regulations establish a specific requirement only for Tartrazine, requiring products containing this additive to declare its full name in the ingredient list, without providing a clear rationale for this distinction ^50^. Together, these differences highlight important variations in how countries address the potential risks associated with food colorings.

While these regulatory measures reflect increasing scientific and policy attention to food colorings, less is known about how parents understand these additives in practice. In the focus groups, although parents understood the function of food colorings, there was some confusion regarding the differences between types, such as natural and synthetic colorings. This distinction appears to be largely technical, as many “natural” colorants are now synthetically produced and may also pose health risks. For instance, Carmine and Annatto have been associated with allergic reactions such as urticaria ^51^. Furthermore, the Brazilian regulatory authority has emphasized that food colorings labeled as “synthetic identical to natural” or “natural” do not offer any advantages in terms of food quality or safety compared to artificial colorants ^51,52^.

Another theme that emerged during the focus groups was concern about the health effects of food colorings, such as allergic reactions, which led some parents to avoid offering products containing these additives to their children. Similar concerns about food additives have been reported in a study conducted in South Korea, where parents of school-aged children identified food additives as a major dietary risk factor. In that study, most parents classified food additives as dangerous or very dangerous to health and believed that foods without additives were healthier ^53^.

Participants also reported difficulty limiting the consumption of products containing food coloring, noting that brightly colored products are particularly attractive to children. Previous studies examining the use of characters as a marketing strategy aimed at children have shown that the presence of characters increases children’s preference for these foods and influences how they perceive their taste, leading them to believe that the products are tastier because of these marketing strategies^54^. Taken together, these findings suggest that the combination of visually appealing product characteristics, such as food coloring, and child-directed marketing strategies may act synergistically to influence children’s food choices and hinder efforts to promote healthy eating in this population. However, further research is needed to specifically assess the combined effect of these factors.

Viewed through the TPB, these findings suggest that limited perceived behavioral control is a key barrier to parents’ intention to avoid food colorings. Parents held negative attitudes toward food colorings and described avoiding them as desirable, but cost, the presence of colorings in most products their children like, and children’s attraction to colorful packages may limit their sense of control. The market analysis shows that these barriers may reflect the food supply, where child-targeted UPFs containing food colorings are widespread (2 in 5 products), with more than 70% of candies and carbonated beverages also containing them. Some purchases may also be responses to the environment, such as product displays and children’s requests in the store, as described by the Prototype/Willingness Model ^17^, which recognizes that behavior may arise not only from reasoned intentions but also from willingness to act in response to environmental opportunities, such as product availability and advertising. Therefore, strategies that rely only on parents’ intentions are therefore unlikely to be enough; changes to the food environment, such as marketing restrictions and clearer labeling, would make the intended behavior easier.

Regarding the identification of food colorings on packaged food labels, most participants correctly identified the beverages with food colorings from the front of the package, but most also judged chocolate milk, which had none, as containing them, which suggests they relied on product type and appearance rather than on label information. In ingredient lists, most participants found the food colorings, but up to half also classified other ingredients, such as flavorings and malt extract, as colorings. Previous research has suggested that information about food additives on packaged food labels is often unclear or insufficient for consumers to interpret accurately ^55^. This lack of transparency compromises consumers’ right to information, posing potential health risks and hindering efforts to promote healthy eating.

These findings support the need for public policies aimed at improving transparency around food additives and reducing children’s exposure to marketing strategies associated with unhealthy products. International organizations have recommended restricting child-directed marketing across multiple settings, including food packaging, schools, and digital media, as well as reducing exposure to additives associated with adverse health effects ^1,56^. Labeling regulations should also ensure clearer disclosure of additives, particularly those potentially harmful to children. Policies adopted in countries such as Mexico and Argentina, which require front-of-package warnings for products containing sweeteners not recommended for children, illustrate potential approaches that could be adapted to other food additives of concern ^57^. Policies that restrict child-directed marketing may also help shift social norms that associate colorful or sweet foods with childhood.

This study has some limitations. The market analysis was restricted to products launched between 2018 and 2021 and may not reflect more recent trends in formulations or marketing practices. In addition, the focus groups included parents from only two municipalities within one Brazilian state, which may limit the transferability of findings to other regions or population groups. Mintel GNPD captures product launches rather than sales, so prevalence estimates are not weighted by consumption, and we assessed the presence, not the amount, of food colorings. Participants were mostly women and college-educated, and food colorings were one topic within a broader discussion on non-sugar sweeteners, which may have limited the depth of discussion, and the questionnaire included only 5 beverages. Finally, although the study explored parents’ perceptions and ability to identify food colorings, it did not assess actual purchasing behaviors or children’s dietary intake.

Despite these limitations, this study has important strengths. The market analysis was based on a large and comprehensive dataset derived from Mintel GNPD and Euromonitor International, ensuring broad coverage of the Brazilian packaged food market. In addition, a validated protocol was used to identify child-targeted marketing strategies, and the qualitative analysis used independent double coding with a structured framework. By integrating market surveillance data with parents’ perceptions, this study contributes important evidence to ongoing discussions about food additive regulation, food labeling policies, and strategies to improve food environments for children.

## 5. IMPLICATIONS FOR RESEARCH, PRACTICE AND POLICY

Food colorings were present in 2 in 5 UPFs with child-targeted marketing in Brazil, and parents, although concerned about these additives, found them hard to avoid and to identify on labels. These findings support restricting child-directed marketing, including packaging, and labeling that makes food colorings easier to identify, alongside nutrition education that helps parents read ingredient lists. Future research could estimate children’s intake of food colorings in Brazil and test whether label formats or education improve parents’ identification of colorings.

## Data Availability

The data used in this study are not publicly available due to the need to protect the privacy of study participants and because food supply data were obtained from a private company. However, metadata may be made available by the corresponding author upon reasonable request.

## Acknowledgments

Access to Mintel-GNPD and Euromonitor International data was provided through a partnership with the Global Food Research Program at the University of North Carolina at Chapel Hill and the Center for Epidemiological Studies in Nutrition and Health at the University of Sao Paulo.

## Funding Acknowledgments

This study was partially funded by Bloomberg Philanthropies through a subaward agreement between the Center for Epidemiological Research in Nutrition and Health (NUPENS), University of São Paulo, and the Global Food Research Program at the University of North Carolina at Chapel Hill. B.S.N. received a master’s scholarship from the Brazilian Coordination for the Improvement of Higher Education Personnel (CAPES) (grant number: 88887.639661/2021-00). M.F.E. received a Young Scientist scholarship from the Brazilian National Council for Scientific and Technological Development (CNPq). M.F.G. received a doctoral fellowship from the Sumner M. Redstone Global Center for Prevention and Wellness at George Washington University. C.Z. is supported by the Chilean National Research and Development Agency – ANID FONDECYT postdoctorado (grant: 3250781).

## Author contributions

B.S.N. and M.F.E.: Writing – review & editing, Writing – original draft, Formal analysis, Data curation, Investigation, Methodology, Conceptualization. M.F.G.: Writing – review & editing, Formal analysis, Investigation, Methodology, Conceptualization. C.Z.: Writing – review & editing, Methodology. A.C.S.: Writing – review & editing, Formal analysis, Methodology, Conceptualization. A.C.D.: Writing – review & editing, Formal analysis, Methodology, Conceptualization, Supervision.

