## Supplementary Material 1, 2, 3, 4, 5 and 6 for "Food Colorings in Child-Targeted Ultra-Processed Foods in Brazil: Market Prevalence and Parental Perceptions"

**Supplementary Materials:**

**Supplementary Material 1.** Descriptions of food colorings, including International Numbering System (INS, Codex Alimentarius) numbers, full names, and similar terms identified among the products.

| **Origin/Group** | **INS** | **Name** | **Similar Terms of *Codex Alimentarius* 2023** | **Similar Terms Identified by the Authors** |
| --- | --- | --- | --- | --- |
| Codex 2023 | 134 | Spirulina Extract |  |  |
| Codex 2023 | 143 | Fast Green Fcf | C.I. Food Green 3; Ci (1975) No. 42053; Fd&C Green No. 3 |  |
| Codex 2023 | 160cii | Paprika Extract |  |  |
| Codex 2023 | 161biii | Lutein Esters From Tagetes Erecta |  |  |
| Codex 2023 | 161hi | Zeaxanthin, Synthetic |  |  |
| Codex 2023 | 183 | Jagua (Genipin-Glycine) Blue |  |  |
| Riboflavin Group | 101i | Riboflavin Synthetic | Riboflavin 5'-Phosphate Ester Monosodium Salt; Vitamin B2 Phosphate Ester Monosodium Salt | Riboflavin |
| Riboflavin Group | 101ii | Riboflavin-5'-Sodium Phosphate' | Vitamin B2 Ester Monosodium Salt |  |
| Riboflavin Group | 101iii | Riboflavin From Bacillus Subtilis |  |  |
| Riboflavin Group | 101iv | Riboflavin From Ashbya Gossypii |  |  |
| Caramel Group | 150a | Caramel I | Caustic Caramel; Plain Caramel | Pure Caramel;Caramel;Caramel Color |
| Caramel Group | 150b | Caramel Ii | Caustic Sulfite Caramel | Sulphite Caramel |
| Caramel Group | 150c | Caramel Iii | Ammonia Caramel | Caramel With Ammonia |
| Caramel Group | 150d | Caramel Iv | Sulfite Ammonia Caramel | Caramel Iv;Caramel With Ammonium Sulfite;Caramel Color Iv |
| Betacarotene Group | 160ai | Beta-Carotene Synthetic | C.I. Food Orange 5 | Beta Carotene Synthetic;Betacarotene Synthetic;Synthetic Beta-Carotene;Synthetic Beta Carotene;Synthetic Betacarotene;Beta Carotene;Betacarotene;160 |
| Betacarotene Group | 160aii | Beta-Carotene Vegetable | Carotenes-Natural; Ci Food Orange 5; Mixed Carotenes; Natural Beta-Carotene | Beta Carotene Vegetable;Betacarotene Vegetable;Vegetable Beta-Carotene;Vegetable Beta Carotene;Vegetable Betacarotene;Vegetable Carotene |
| Betacarotene Group | 160aiii | Blakeslea Trispora-Beta-Carotene |  | Blakeslea Trispora-Beta Carotene;Blakeslea Trispora-Betacarotene |
| Betacarotene Group | 160aiv | Β-Carotene-Rich Extract From Dunaliella Salina |  | Dunaliella Salina Extract Rich In Beta-Carotene;Dunaliella Salina Extract Rich In Beta Carotene |
| Betacarotene Group | 160e | Carotenal, Beta-Apo-8'- | C.I. Food Orange 6 | Beta-Apo-8'-Carotenal |
| Anatto | 160bi | Annatto Extracts, Bixin-Based |  | Bixin;Anatto;Anato;Annato;160b |
| Anatto | 160bii | Annatto Extracts, Norbixin-Based |  | Norbixin |
| Lycopene Group | 160di | Lycopene Synthetic |  | Synthetic Lycopene;Lycopene;160d |
| Lycopene Group | 160dii | Lycopene Tomato |  | Tomato Lycopene |
| Lycopene Group | 160diii | Lycopene, Blakeslea Trispora |  | Blakeslea Trispora Lycopene |
| Iron Group | 172i | Iron Oxide, Black | C.I. Pigment Black 11; Ci (1975) No. 77499 | Black Iron Oxide;172 |
| Iron Group | 172ii | Iron Oxide, Red | C.I. Pigment Red 101; C.I. Pigment Red 102; Ci (1975) No. 77491 | Red Iron Oxide |
| Iron Group | 172iii | Iron Oxide, Yellow | C.I. Pigment Yellow 42; C.I. Pigment Yellow 43; Ci (1975) No. 77492 | Yellow Iron Oxide |
| Codex 2023 | 181 | Tannic Acid | Gallotannic Acid; Tannins (Food Grade) | Tannins;Tannin |
| Codex 2023 | 123 | Amarant | Ci (1975) No. 16185;  Ci Food Red 9;  Naphtol Rot S. |  |
| Codex 2023 | 104 | Quinoline Yellow | Ci (1975) No. 47005;  Ci Food Yellow 13 | Yellow Quinoline |
| Codex 2023 | 110 | Sunset Yellow | Ci (1975) No. 15985;  Ci Food Yellow 3;  Crelborange S;  Fd&C Yellow No. 6 | Yellow Sunset;Sunset Yellow Fcf |
| Codex 2023 | 122 | Azorubin | Carmoisine;  Ci (1975) No. 14720;  Ci Food Red 3 | Carmoisin |
| Codex 2023 | 133 | Brilliant Blue Fcf | Ci (1975) No. 42900; Ci Food Blue 2; Fd&C Blue No.1 | Brilliant Blue |
| Codex 2023 | 161g | Canthaxanthin | Ci (1975) No 40850; Ci Food Orange 8 |  |
| Codex 2023 | 170i | Calcium Carbonate | Chalk | 170 |
| Codex 2023 | 120 | Carmine | Carmine; Ci (1975) No. 75470;  Ci Natural Red 4;  Cochineal Carmine | Carmin |
| Codex 2023 | 140 | Chlorophylls | C.I. (1975) No. 75810; Ci Natural Green 3; Magnesium Chlorophyll; Magnesium Phaeophytin | Chlorophyl |
| Codex 2023 | 141i | Chlorophylls-Copper Complex | C.I. (1975) No. 75810; Ci Natural Green 3; Copper Chlorophyll; Copper Phaeophytin | Chlorophylls Copper Complex |
| Codex 2023 | 141ii | Chlorophyllines-Copper Complexes, Potassium And Sodium Salts | C.I. (1975) No. 75810; Potassium Copper Chlorophyllin; Sodium Copper Chlorophyllin | Chlorophyllines Copper Complexes, Potassium And Sodium Salts |
| Codex 2023 | 100i | Curcumin | C.I. Natural Yellow 3; Diferuloymethane; Kurkum; Turmeric Yellow |  |
| Codex 2023 | 171 | Titanium Dioxide | Ci (1975) No. 77891; Ci Pigment White 6 | Dioxide Titanium |
| Codex 2023 | 127 | Erythrosine | C.I. (1975) No. 45430;  C.I. Food Red 14;  Fd&C Red No. 3 |  |
| Codex 2023 | 163ii | Grape Skin Extract | Eno; Enociania |  |
| Codex 2023 | 132 | Indigotine | C.I. Food Blue 1; Ci (1975) No. 73015; Fd&C Blue No. 2; Indigo Carmine | Indigo Carmine;Indigo |
| Codex 2023 | 161bi | Lutein From Tagetes Erecta |  | Tagetes Erecta Lutein |
| Codex 2023 | 155 | Brown Ht | Chocolate Brown Ht; Ci (1975) No. 20285; Ci Food Brown 3 |  |
| Codex 2023 | 151 | Glossy Black | Black Bn; Black Pn; Brilliant Black Bn; Ci (1975) No. 28440; Ci Food Black 1 | Black Pn;Brilliant Black |
| Codex 2023 | 124 | Ponceau 4r | Ci (1975) No. 16255;  Ci Food Red 7;  Cochineal Red A;  New Coccine | Ponceau;Cochinilla |
| Codex 2023 | 129 | Red Allura | Ci (1975) No.16035; Ci Food Red 17; Fd&C Red No.40 | Red Alura;Red 40;Red N40;Red No. 40 |
| Codex 2023 | 162 | Purée Red | Beetroot Red | Red Purée |
| Codex 2023 | 102 | Tartrazine | Ci (1975) No. 19140;  Ci Food Yellow 4;  Fd&C Yellow No. 5 |  |

**Supplementary Material 2.** Focus group sociodemographic questionnaire.

| Are you a parent or caregiver of a child (ages 2–11) currently living with you? | 1=Yes  2=No |
| --- | --- |
| In the last month, how much of the household's food shopping did you do? | 1=None  2= Less than half  3=Half  4=More than half  5=All |
| What is your relationship to the child? | 1=Mother  2=Father  3=Other (specify: ____________) |
| Does the child live with their parents? | 1=Yes, with both  2=Yes, only with the mother  3=Yes, only with the father  4=No, other (specify: ____________) |
| What is your age (in years)? |  |
| How would you describe your gender identity? | 1=Male  2=Female  3=Other |
| How do you identify in terms of race or ethnicity? | 1=Brown  2=Black  3=White  4=Asian  5=Other |
| What is the total monthly income of everyone living in your household? | 1=Up to R$ 2.200  2=R$ 2.200 to R$ 5.500  3= R$ 5.500 to R$ 11.000  4= R$ 11.000 to R$ 22.000  5=Above R$ 22.000 |
| What is the highest level of education you have completed? | 1=None  2=Elementary School  3= Upper Elementary  4= High School  5= Trade school 6 = Undergraduate degree  7=Graduate degree |
| What is your marital status? | 1= Single  2= Married or living with a partner  3= Divorced or separated  4= Widowed |
| Including the child, how many people live in your household? | _____ People |
| Of those, how many are under 10 years old? | ______chidren |
| What is your current work schedule outside the home? | 1=0 hours  2=0 to 20 hours per week  3=21 to 40 hours per week  4= More than 40 hours per week |
| What is the age of the child? (If you have more than one child within the study’s age range, please provide the age of the oldest one.) |  |

**Supplementary Material 3.** Survey to assess parents’ ability to identify food colorings on product labels.

1. Below are several examples of different products. We want to explore your understanding of beverages sold in the market.
   a. Please circle which of these products you are familiar with (consider these brands or similar ones).

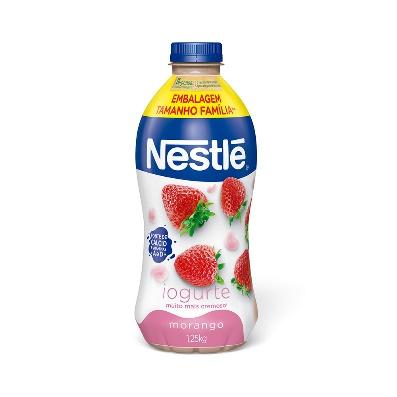

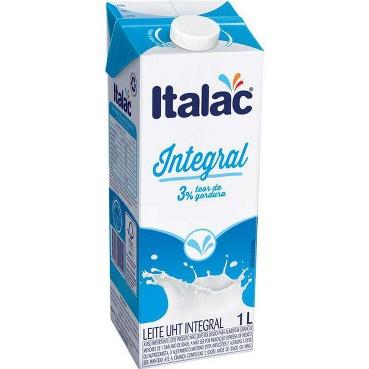

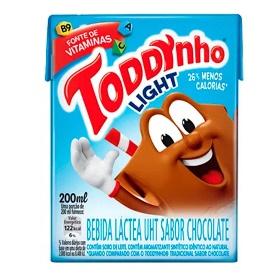

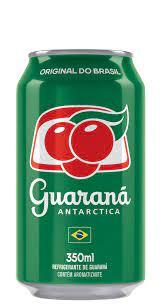

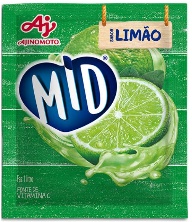

b. Please circle which of these products you purchase for the child (consider these brands or similar ones).

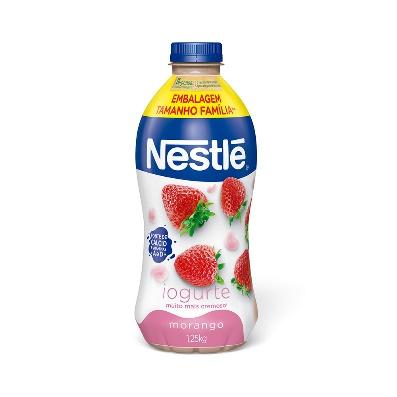

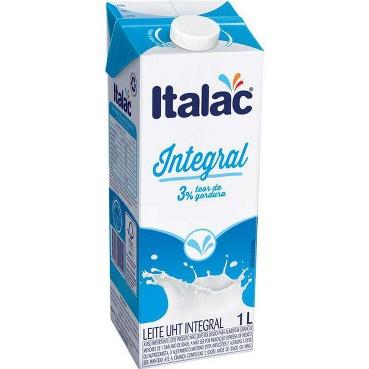

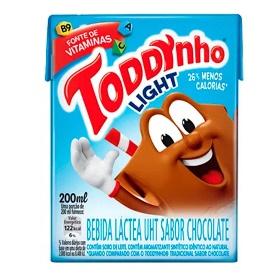

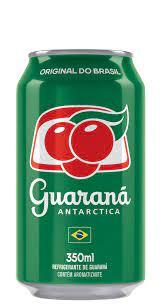

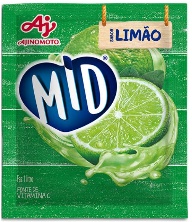

c. Now, ONLY REGARDING THE PRODUCTS AND BRANDS LISTED BELOW.

Please circle which of these products you think contain COLORINGS.

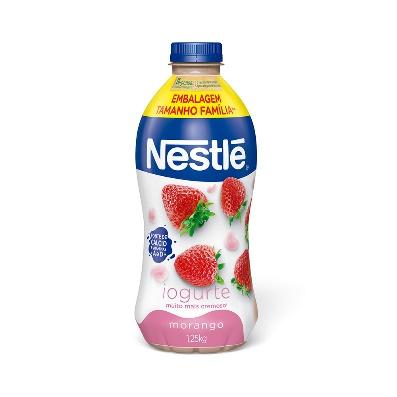

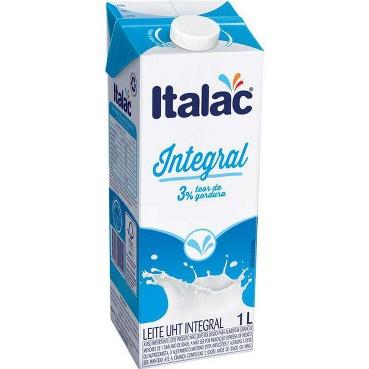

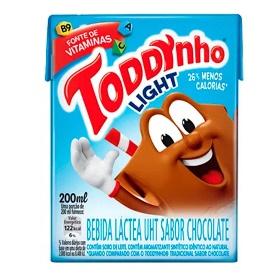

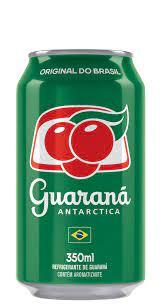

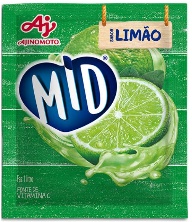

2. Still about FOOD COLORINGS.

Below are some examples of ingredient lists that you probably have seen on the back of beverage packaging. Please circle which ingredients you believe are non-sugar sweeteners.

i. Ingredients: Partially skimmed fermented milk and/or whole pasteurized milk, strawberry preparation (water, sugar syrup, strawberry pulp, modified starch, sugar, tricalcium phosphate, flavorings, citric acid acidulant, guar gum thickener, natural carmine colorant, potassium sorbate preservative), whey and/or reconstituted whey, sugar syrup, whey powder, and lactic cultures.

ii. Ingredients: Whole milk and stabilizers (sodium citrate, sodium triphosphate, monosodium monophosphate, and disodium diphosphate).

iii. Ingredients: Reconstituted skimmed milk, hydrated cocoa, reconstituted whey, reconstituted whole milk, sugar, malt extract, vitamins (C, A, and folic acid), thickeners (gellan gum, sodium carboxymethyl cellulose, and guar gum), sodium citrate stabilizer, sweeteners (acesulfame potassium and sucralose), soy lecithin emulsifier, and flavoring.

iv. Ingredients: Carbonated water, sugar, guaraná extract, citric acid acidulant, preservatives (sodium benzoate and potassium sorbate), flavoring, and caramel color.

v. Ingredients: Sugar, dehydrated lemon juice, maltodextrin, vitamin C (ascorbic acid), citric acid acidulant, natural lemon flavoring, tricalcium phosphate anticaking agent, artificial sweeteners (per 100 mL: aspartame 24.6 mg, acesulfame potassium 10.1 mg, and advantame 0.01 mg), acidity regulator (sodium citrate), inorganic colorant (titanium dioxide), and artificial colorant (tartrazine).

**Supplementary Material 4.** Semi-structured focus group discussion guide.

1. What type of beverage do you give to the child?
2. What factors influence the types of beverages you offer to the child?
3. What characteristics do you look for in beverages when choosing what to give to the child? (nutrients, ingredients)
   a) Are there certain nutrients/ingredients you want to provide to the child? Why or why not?
   b) Are there certain nutrients/ingredients you try to avoid for the child? Why or why not?
4. What do you understand by ‘coloring’ in beverages?
5. What is your opinion on colorings in beverages?
   a) What do you think about giving colorings to the child?
      i. How much do you see them as healthy or unhealthy for the child to consume?

**Supplementary Material 5.** Sociodemographic characteristics of focus group participants.

|  | N | % |
| --- | --- | --- |
| *Parents’ sex* |  |  |
| Male | 5 | 12.5 |
| Female | 35 | 87.5 |
| *Parents’ race* |  |  |
| Black | 16 | 40.0 |
| White | 24 | 60.0 |
| *Educational attainment* |  |  |
| Upper Elementary | 1 | 2.5 |
| High School | 15 | 37.5 |
| Trade school | 2 | 5.0 |
| Undergraduate degree | 8 | 20.0 |
| Graduate degree | 14 | 35.0 |
| *Child’s school* |  |  |
| Private | 13 | 32.5 |
| Public | 27 | 67.5 |

**Supplementary Material 6.** Description of participants’ familiarity with and purchase of the beverages presented in the survey.

|  | Flavored yogurt | Milk | Chocolate-flavored milk | Soda | Powdered juice |
| --- | --- | --- | --- | --- | --- |
|  | % | % | % | % | % |
| Familiar with the product | 100 | 100 | 100 | 97,4 | 89,7 |
| Usually purchase the product | 56,4 | 61,5 | 30,7 | 35,9 | 38,4 |
